# Capturing heterogeneity in *Opisthorchis viverrini* epidemiology and control

**DOI:** 10.1101/2023.05.09.23289707

**Authors:** Lars Kamber, Christine Bürli, Helmut Harbrecht, Peter Odermatt, Somphou Sayasone, Nakul Chitnis

## Abstract

*Opisthorchis viverrini* is a parasitic liver fluke affecting over 10 million people despite sustained control efforts. High intensity infections are a risk factor for the often fatal bile duct cancer, cholangiocarcinoma. Similar to other helminthiases, the distribution of worm burden in humans is highly uneven within populations. We developed multiple models which allow us to capture heterogeneity in transmission and interventions dynamics and the resulting impact on worm distribution: An agent-based model with the common assumption of gamma-distributed transmission parameters; an agent-based model with an alternative nonparametric distribution of transmission parameters; and a simpler ordinary differential equation model. We calibrated all models to prevalence and intensity of infection data in humans, and prevalence data for reservoir hosts and intermediate hosts from southern Lao People’s Democratic Republic. We simulated the impact of multiple interventions on prevalence, intensity of infection and the distribution of worm burden in humans. Our results showed significant overlap in predictions of prevalence and intensity of infection over time between the agent-based models and the ordinary differential equation model, corroborating both the simple and more complex models; however, the nonparametric model was better able to capture the distribution of the highest intensity burden in individuals. Under assumptions of homogeneous adherence to mass drug administration campaigns, no model was able to capture the changing heterogeneity of worm burden over time seen in the epidemiological data. Allowing for heterogeneous adherence in these campaigns, which was only possible in the agent-based models, allowed us to explain the changes seen in the worm distribution and burden seen in the data. This result highlights the added benefit of agent-based models in capturing the changing heterogeneity in worm burden in areas with repeated mass treatments. Appropriately capturing this heterogeneity is essential in understanding the relationship between worm burden, control interventions and subsequent disease burden.

**Author summary:** *Opisthorchis viverrini* is a parasitic liver fluke affecting over 10 million people despite sustained control efforts. The distribution of worm burden in humans is highly uneven within populations with high intensity infections being a major risk factor for bile duct cancer. We developed and present multiple models, some of which allow us to capture this uneven distribution in susceptibility to infection as well as in adherence to treatment: Two agent-based models of high complexity and a simpler population-based model. We calibrated all models to replicate worm burden data collected in southern Lao People’s Democratic Republic. We simulated the impact of multiple interventions and showed significant overlap of all models in many aspects, corroborating both the simple and more complex models. However, we show that the agent-based models have the added benefit of being able to better capture the unevenness of worm burden before and especially after interventions.

## 1 Introduction

*Opisthorchis viverrini* is a parasitic liver fluke endemic to the Greater Mekong Subregion (GMS) in Southeast Asia [1]. While substantial reductions in *O. viverrini* prevalence were achieved over recent decades, a total of 12.4 million individuals in the GMS were estimated to still be infected in 2018 [2–4]. Infections with *O. viverrini* can be asymptomatic [5] but frequently cause liver morbidity and, especially after sustained heavy infection, cholangiocarcinoma, a malignancy with poor prognosis [6–9].

Humans and other mammals, such as dogs and cats, serve as definitive hosts of *O. viverrini* [10]. The mature parasite resides in the bile duct producing eggs which reach the environment with the hosts’ stool. *Miracidia* hatch from eggs and are ingested by freshwater snails of the genus *Bithynia*, inside which they develop into *cercariae*. The *cercariae* shed from the snails, penetrate the skin of fish from the carp family and develop into *metacercariae* inside the fish. When definitive hosts consume infected fish in raw or undercooked form, the *metacercariae* migrate to the hosts’ bile duct and develop into adult worms that can live for many years.

The intensity of liver fluke infection is highly heterogeneous in populations where *O. viverrini* is endemic, with relatively few individuals exhibiting very heavy worm burden [11]. This pattern of aggregation is typical for macroparasitic infections and has been explained by differences in exposure, susceptibility and immune response of individuals [12, 13].

Praziquantel is the drug of choice for the treatment of *O. viverrini* infection. Non-pharmaceutical interventions include education campaigns on safe fish consumption and use of sanitation, improvement of sanitation, snail control and safe fish farming. Mass drug administration (MDA) campaigns with Praziquantel are regularly conducted in endemic areas, but with limited success [14]. While the drug itself shows high efficacy when dosed correctly, infection prevails because of limited coverage, limited adherence and continued consumption of raw and undercooked fish.

In previous work, we developed various population-based models (PBMs) for *O. viverrini* transmission and control. These are ordinary and partial differential equation models that track the mean worm burden in humans and reservoir hosts, and the prevalence of infection in intermediate hosts. The main conclusions from PBM studies are that interruption of transmission can be achieved by only targeting humans with interventions [15], but MDA campaigns covering all age groups are necessary to interrupt transmission and treating school-aged children exclusively is not sufficient to achieve this goal [16]. Other modelling work on *O. viverrini* has focused on the effect of ecological and climate factors on transmission [17, 18]. Furthermore, several compartmental models for the transmission and control of *Clonorchis sinensis*, a parasite with the same life cycle as *O. viverrini*, have been published [19–23].

While PBMs can provide distributions of worm burden as an output based on mean worm burden [12], they usually assume homogeneity of individuals when it comes to susceptibility to infection, behavior, adherence to interventions and coverage of interventions. PBMs can accommodate for heterogeneity to some degree by adding compartments for each combination of considered characteristics. For example, separate compartments were added for individuals with different propensity to consume undercooked fish in a PBM for *C. sinensis* [21]. However, when modelling multiple sources of heterogeneity simultaneously, the number of required compartments grows exponentially. Agent-based models (ABMs) track each individual separately, allowing heterogenous characteristics, and tracking parasite and disease burden at the individual level. Because heterogeneities among humans play an important role in the transmission, pathology and treatment of *O. viverrini*, we have developed an ABM for *O. viverrini* transmission and control. To our knowledge, this is the first model of this kind for *O. viverrini* or *C. sinensis*. We present two versions of the ABM: One with the assumption of gamma-distributed transmission parameters over the population, which is the most common assumption in ABMs of schistosomiasis [24], and one with a nonparametric distribution of transmission parameters derived from the distribution of worm burden in two villages in Lao PDR. We fit both models to this data and compare the equilibrium state and intervention impact predictions to those of a previously published PBM of *O. viverrini* transmission and control [25]. We show that taking heterogeneities into account is essential to adequately model the impact of interventions on worm distribution.

## 2 Methods

### 2.1 Data

We use previously published data collected in cross-sectional surveys on two adjacent Mekong river islands in Southern Laos in 2012 and 2018 [11]. Between the surveys, MDA was conducted every year except in 2013 and education campaigns were carried out. The 2012 data includes the intensity of infection for *O. viverrini* measured in eggs per gram of stool (EPG) in stool samples of humans, cats and dogs. It also includes prevalence of infection in the intermediate snail and fish hosts. For the year 2018, only EPG data in humans is available. The number of samples of each kind are listed in Table 1 and the distribution of EPG among humans is shown in Fig 1. We transform EPG to worm counts and vice versa in individuals and animals with a density-dependent function derived from a purging study as described in the supplementary Section S1.6.2.

**Table 1.** Number of tested and positive samples from definitive and intermediate hosts in 2012 and 2018.

|  | 2012 |  |  |  |  | 2018 |
| --- | --- | --- | --- | --- | --- | --- |
|  | Humans | Cats | Dogs | Snails | Fish | Humans |
| Tested | 994 | 64 | 68 | 3102 | 628 | 580 |
| Positive | 603 | 34 | 17 | 9 | 169 | 135 |
| Prevalence | 60.7% | 53.1% | 25% | 0.3% | 26.9% | 23.3% |

**Fig 1.**
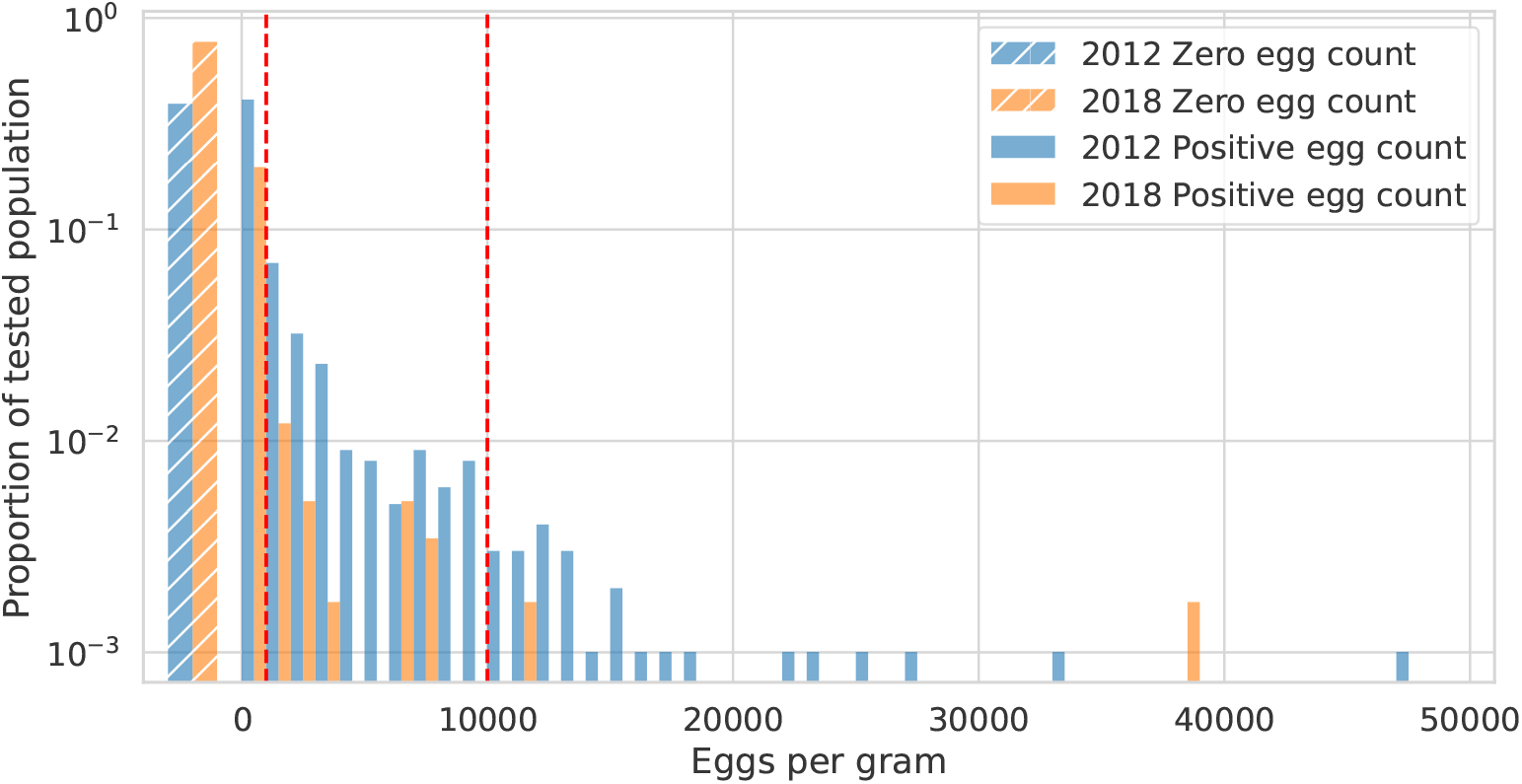
Distribution of EPG among humans in data collected in 2012 and 2018. The hatched wide bars on the left show the proportion of individuals with an egg count of zero. The remaining bars show the proportion of individuals with nonzero egg counts, where the bin width equals 1000 EPG. The red lines mark the thresholds of World Health Organization worm burden classification with 1-999 EPG being classified as light infection, 1000-9999 as moderate infection and 10000 and above as heavy infection [26]. The EPG data from both years show a high degree of aggregation, *k* = 0.10 in 2012 and *k* = 0.03 in 2018, implying more aggregation in 2018. These values of *k* are obtained by applying Equation (3) to the EPG data.

### 2.2 Agent-based Model

This section provides a high-level overview of the ABM. A detailed description written according to the ODD (Overview, Design concepts, Details) protocol for describing individual- and agent-based models [27, 28] is provided in the supplementary Section S1. A schematic of the ABM is provided in Fig 2. State variables of *N* humans are tracked individually while animals are modelled at the aggregate level (Table 2). The transmission dynamics are governed by the parameters listed in Table 3. We introduce heterogeneity by letting the worm acquiring rate *β*_*hf,i*_ vary between individuals, *i*. We consider two distributions for randomly drawing *β*_*hf,i*_ and refer to the models using these distributions with separate names: The ABM^Γ^ assumes a gamma distribution, which can be considered the standard assumption for macroparasite models [24]; the AMB^*ε*^ assumes a nonparametric distribution derived from the 2012 field data. The gamma distribution in the ABM^Γ^ is parameterized by the mean and variance parameters 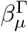 and 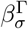 whose values are determined by model fitting. In the AMB^*ε*^, a worm count 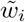 is drawn for each individual from the distribution of worm counts in the 2012 data. The individuals’ *β*_*hf,i*_ is derived from the drawn 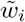 using the relation

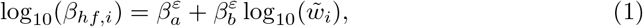

for nonzero values of 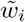, where 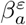 and 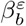 are parameters that are determined by model fitting. If 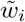 equals zero, *β*_*hf,i*_ is set to zero as well.

**Table 2.** State variables in the ABM. All variables with superscript (*t*) can change over time. The upper part of the table contains the variables which are tracked for each individual human as indicated by subscript *i*. The lower part of the table contains the state variables of nonhuman hosts. The maximum number of worms an individual can carry is set to a biologically implausible value to facilitate the parameter fitting process and is never reached under fitted parameter values.

| Variable | Type | Description | Range |
| --- | --- | --- | --- |
| $\text{sex}_i$ | Boolean | 0 for female, 1 for male | $\{0, 1\}$ |
| $\text{age}_i^{(t)}$ | Float | Age in days | $[0, 100 \times 365]$ |
| $\text{worms}_i^{(t)}$ | Integer | Number of adult worms in individual | $[0, 10^5]$ |
| $\text{epg}_i^{(t)}$ | Integer | Eggs per gram in individual's stool | $[0, \rho(10^5)]$ |
| $\text{beta\_multiplier}_i^{(t)}$ | Float | Multiplier to $\beta_{hf,i}$ from education campaign | $[0, 1]$ |
| $\text{eating}_i^{(t)}$ | Boolean | Individual is consuming under-cooked fish | $\{0, 1\}$ |
| $\text{latrine}_i^{(t)}$ | Boolean | Individual has access to a latrine | $\{0, 1\}$ |
| $\text{latrine\_use}_i^{(t)}$ | Boolean | Individual uses latrine if accessible | $\{0, 1\}$ |
| $\text{dogs}^{(t)}$ | Integer | Total number of worms in dogs | $\geq 0$ |
| $\text{cats}^{(t)}$ | Integer | Total number of worms in cats | $\geq 0$ |
| $\text{snails}^{(t)}$ | Integer | Number of infected snails | $[0, 10^7]$ |
| $\text{fish}^{(t)}$ | Integer | Number of infected fish | $[0, 10^6]$ |

**Table 3.** Transmission parameters of the ABM. Further parameters of the ABM are described in the supplementary Section S1.6.

| Parameter | Description | Unit |
| --- | --- | --- |
| $\beta_{sh}$ | Transmission rate to snails from eggs in human stool | 1/(Day × Animals) |
| $\beta_{sd}$ | Transmission rate to snails from eggs in dog stool | 1/(Day × Animals) |
| $\beta_{sc}$ | Transmission rate to snails from eggs in cat stool | 1/(Day × Animals) |
| $\beta_{fs}$ | Transmission rate to fish from snails | 1/(Day × Animals) |
| $\beta_{hf,i}$ | Transmission rate to individual $i$ from fish | 1/(Day × Animals) |
| $\beta_{df}$ | Transmission rate to dogs from fish | 1/(Day × Animals) |
| $\beta_{cf}$ | Transmission rate to cats from fish | 1/(Day × Animals) |

**Fig 2.**
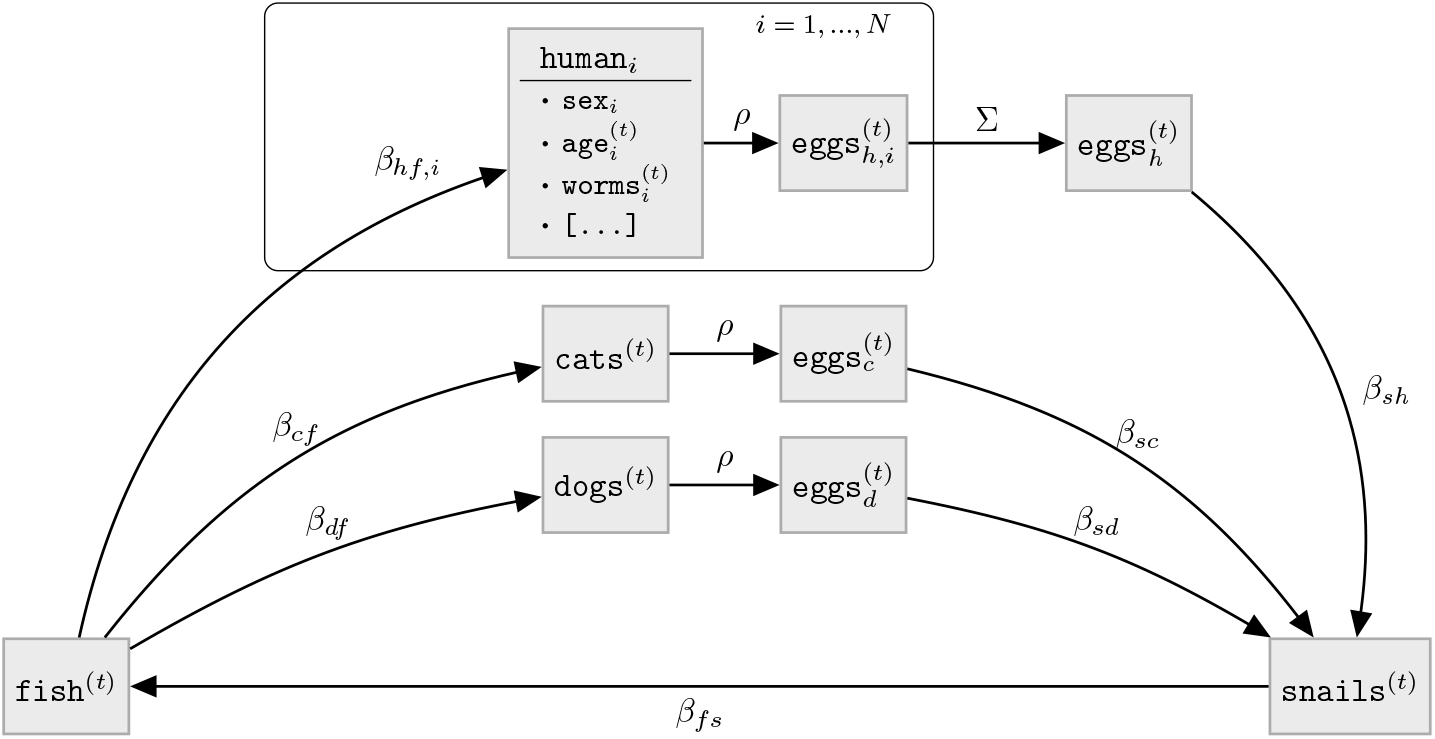
Schematic of the the agent-based model for *O. viverrini* transmission and control. The state variables are described in Table 2 and the transmission parameters are described in Table 3. Worm counts are mapped to egg output with a nonlinear monotonically increasing piecewise function *ρ* derived from purging studies (Section S1.6.2). While *ρ* is applied to the mean worm burden in cats and dogs and then multiplied with the number of cats and dogs, respectively, it is applied to each individual worm count in humans and subsequentially summed up over humans.

For all humans and animals the uptake of new parasites is Poisson distributed with a rate determined by the current state of the system, the transmission parameters, the time step size and the population sizes. We choose a time step size of one month and a population size of 100,000 for fitting and the analyses in this paper.

### 2.3 Population-based model

The PBM we use for comparison with the ABM is an ordinary differential equation model that has been described and analysed in previous work [15, 25]. A schematic of the model is given in Fig 3. The equations and a complete table of parameters are given in the supplementary Section S2.

**Fig 3.**
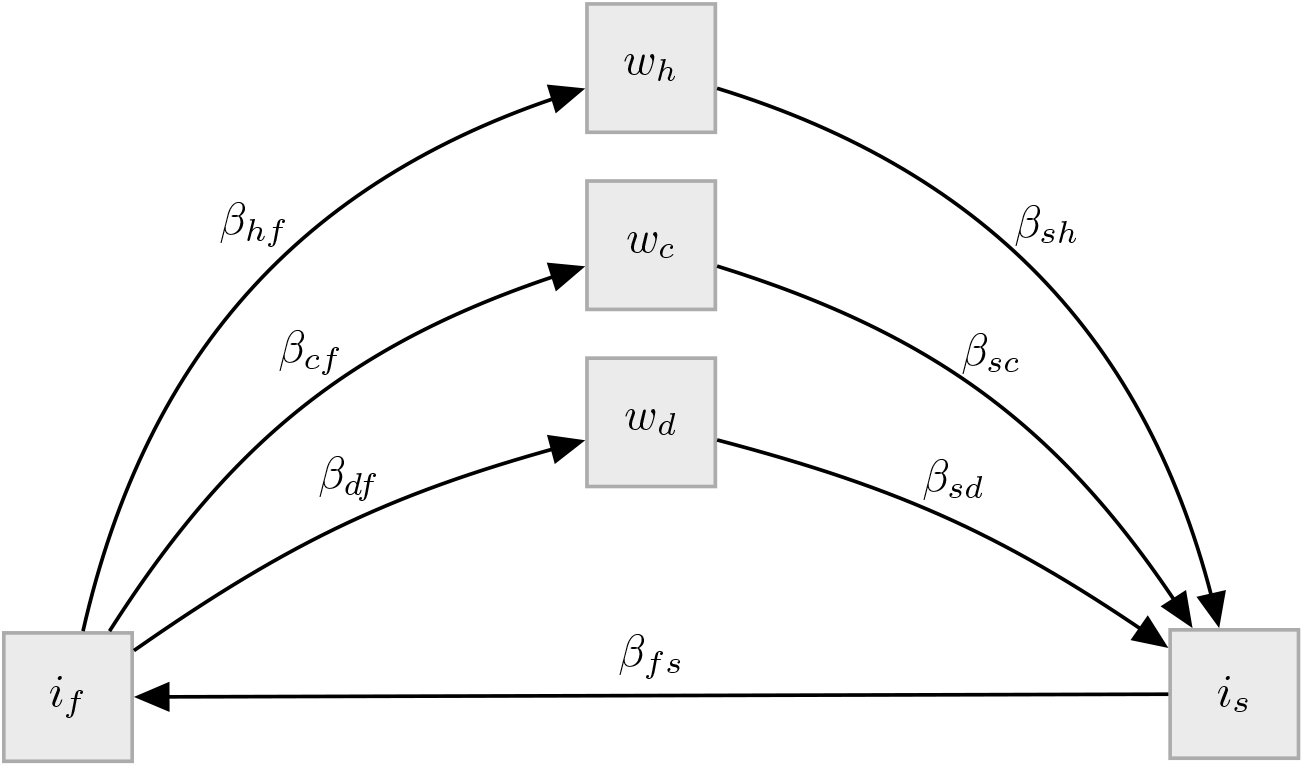
Schematic of the population-based model for *O. viverrini* transmission and control. The state variables are described in Table 4, the transmission parameters roughly correspond to the transmission parameters of the ABM listed in Table 3. As opposed to the ABM, where *β*_*hf,i*_ varies over individuals, there is only a single *β*_*hf*_ in the PBM. Also, the PBM does not translate worm burden in definitive hosts to egg output which results the in the transmission parameters *β*_*sh*_, *β*_*sd*_ and *β*_*sc*_ acting on mean worm burden instead of egg count. Further details of the PBM are provided in supplementary Section S2.

Instead of tracking worms in humans individually, the PBM tracks the mean number of worms across humans. This means that the PBM, as opposed to the ABM, does not provide a distribution of worm burden and therefore prevalence as a direct output. To obtain a distribution, we use a negative binomial parameterized by an aggregation parameter *k* having the probability mass function,

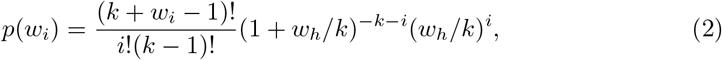

which provides the distribution of worm counts, *w*_*i*_, for a given mean worm burden, *w*_*h*_ [12]. The prevalence, *P*, which corresponds to the proportion of non-zero worm counts, is given by

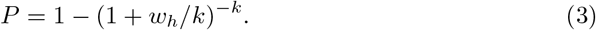

**Table 4.** State variables of the population-based model.

| Variable | Description |
| --- | --- |
| $w_h$ | Mean worm burden per human host |
| $w_d$ | Mean worm burden per dog host |
| $w_c$ | Mean worm burden per cat host |
| $i_s$ | Proportion of infectious snails |
| $i_f$ | Proportion of infectious fish |

We substitute the mean worm burden and prevalence from the field data collected in 2012 for *w*_*h*_ and *P* in Equation (3) and solve numerically for *k* to estimate the level of aggregation in the field data. We then assume *k* to be constant over time [12] and substitute the calculated value of *k* into Equation (2) to obtain a distribution of individual worm burden, *w*_*i*_, for any mean worm burden, *w*_*h*_, calculated during a PBM model run.

### 2.4 Transmission parameter fitting

We fit the transmission parameters included in Table 3 to the 2012 data. The data from 2018 is not used to fit transmission parameters. For both ABMs, we fit the parameters which govern the distribution of *β*_*hf,i*_ as described in Section 2.2. The parameters *β*_*sh*_, *β*_*sc*_ and *β*_*sd*_ are not identifiable for all models given the data and we fix these three parameters to be equal to a parameter *β*_*sx*_ which is fitted instead.

The transmission parameters are calibrated such that the models reach an equilibrium that matches various target summary statistics calculated from the 2012 data listed in Table 5. To create summary statistics that capture the worm distribution in a clinically significant way, we bin the worm burden of positive humans into 3 categories according to World Health Organization criteria [26]: Individuals having between 1 EPG and 999 EPG are classified as having low worm burden, between 1,000 EPG and 9,999 EPG as medium worm burden and above 10,000 EPG as high worm burden. The summary statistics used for fitting are the proportions of individuals that have low, medium or high worm burden as well as the mean of EPG within each of those groups. We use these distributional summary statistics only for the fitting of the ABM^*ε*^, as the ABM^Γ^ and the PBM cannot reach the target prevalence when simultaneously being fitted to the distributional target values. In contrast, the distribution of *β*_*hf,i*_ in the ABM^*ε*^ has a proportion of individuals with a value of zero that results in the target prevalence by design.

**Table 5.**
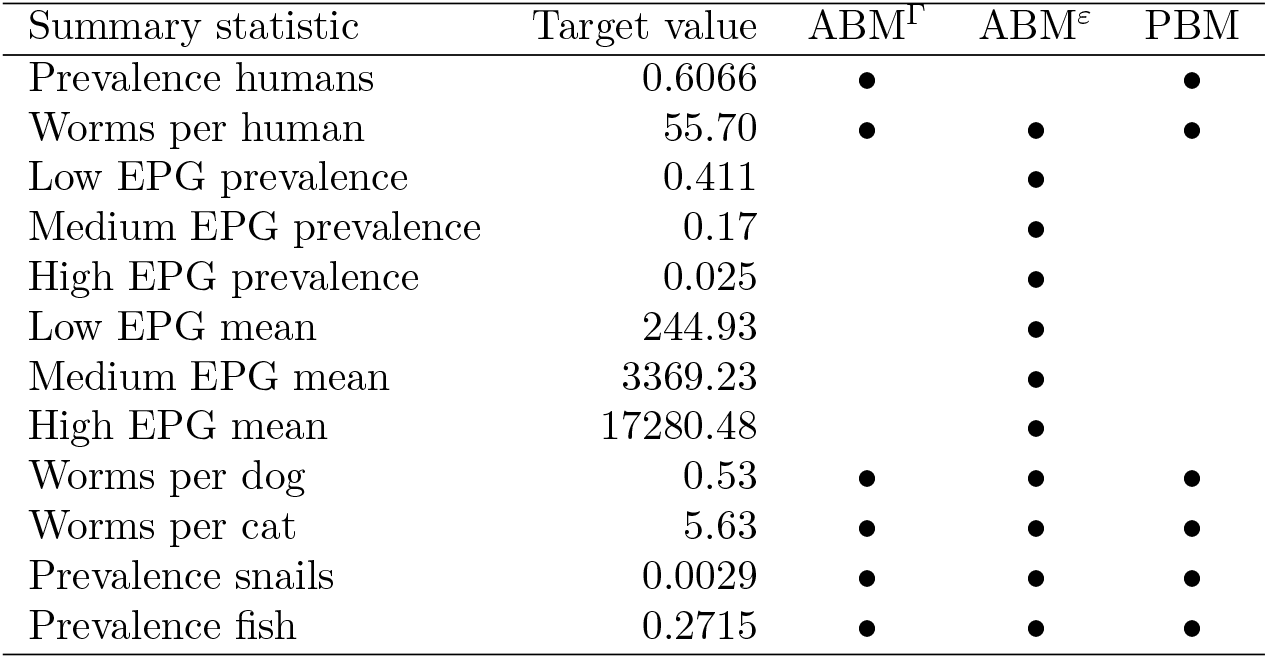
Target summary statistic values used to fit transmission parameters. A bullet signifies that the summary statistic was used to fit the respective model.

We fit the ABMs using Trust Region Bayesian Optimization (TuRBO) with local Gaussian processes implemented with the Python package BoTorch [29, 30]. The PBM parameters required to reach a target equilibrium were calculated analytically.

### 2.5 Mass drug administration modelling

For the comparison of intervention impact predictions, we start all models at equilibrium, run them for one year without interventions and subsequently deploy yearly MDA for five years with coverages constant over time ranging from 0% to 90%. In the ABMs, this corresponds to reducing the worm count to 0 for a random selection of individuals corresponding to the coverage. For the PBM model, we reduce the mean worm count by the MDA coverage. We run each ABM simulation with 10 random seeds. To compare the predictions over different coverages we assess the state variables one year after the last round of MDA. This scheme of five years of MDA and subsequent assessment approximates the MDA and survey scheme of the population from which the data in 2012 and 2018 was obtained (Section 2.1).

### 2.6 Heterogeneity in access to MDA and education campaign

In order to match the model predictions for prevalence and mean worm burden to the 2018 data in a second step, we employ heterogeneity in MDA adherence and include an education campaign. For modelling heterogeneity in MDA adherence, treatment is still distributed randomly but individuals have different probabilities of being reached by MDA campaigns which stay fixed over time. We assume these probabilities to follow a beta distribution with mean 0.8 and examine a range of variances between 0 and 0.3, where a larger variance implies increased heterogeneity. The mean of 0.8 translates to a coverage of 80%, which corresponds to the estimated effective coverage in the study area between 2012 and 2018 as estimated by expert opinion. We assign the drawn probabilities of MDA participation to individuals in descending order of their *β*_*hf,i*_. This means that those individuals with the highest rate of acquiring worms *β*_*hf,i*_ are least likely to be reached by the MDA campaign. As the individuals’ probabilities of being reached during the MDA campaigns stays fixed over time, this also implies serial correlation of MDA adherence.

For the yearly education campaign, we assume that all individuals are reached and that, for each campaign, the effect is the multiplication of 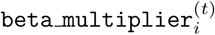 with a factor equal to one minus the education campaign’s efficacy, which we examine within a range of 0 to 0.5.

We run simulations for one year in equilibrium followed by five years of yearly MDA and education campaigns and assess the state variables one year after the final intervention. The simulations are run over combinations within the explored range of MDA variance and education campaign efficacy. Using Gaussian process regression, we predict the combinations of education campaign efficacy and MDA variance which reproduce the mean worm burden and prevalence from the 2018 data at the end of the model run.

## 3 Results

We obtain good fits in equilibrium for all the the target summary statistics used for fitting with the parameter values listed in Table 6. A kernel density estimate of the worm distribution among the individuals in the 2012 data and in the models at equilibrium is provided in Fig 4. It shows a very high agreement between the ABM^*ε*^ and the data, while the equilibrium state of the PBM and the ABM^Γ^ are very similar but show some discrepancy to the data. This high agreement in equilibrium worm distribution between ABM^Γ^ and the PBM can be expected, since we assume that the worm uptake of individuals in the ABMs follows a Poisson distribution at each time step.

**Table 6.** Fitted parameter values for all models. The first five parameters in the table are parameters determining the transmission from snails to humans and are specific to the respective models. The remaining parameters are part of all three models. The fitted values for *β*_*df*_, *β*_*cf*_ and *β*_*fs*_ are very close to each other for all models. The fitted values for *β*_*sx*_ are close for the ABMs but different for the PBM since *β*_*sx*_ acts on the total egg output in the ABMs but acts on mean worm burden in the PBM.

| Parameter | $ABM^\Gamma$ | $ABM^\epsilon$ | PBM |
| --- | --- | --- | --- |
| $\log_{10} \beta_\mu^\Gamma$ | -7.14 | – | – |
| $\log_{10} \beta_\sigma^\Gamma$ | -13.45 | – | – |
| $\beta_a^\epsilon$ | – | -8.88 | – |
| $\beta_b^\epsilon$ | – | 0.99 | – |
| $\log_{10} \beta_{hf}$ | – | – | -7.25 |
| $\log_{10} \beta_{df}$ | -8.88 | -8.80 | -8.88 |
| $\log_{10} \beta_{cf}$ | -7.84 | -7.85 | -7.84 |
| $\log_{10} \beta_{sx}$ | -15.96 | -15.83 | -11.85 |
| $\log_{10} \beta_{fs}$ | -7.87 | -7.86 | -7.87 |

**Fig 4.**
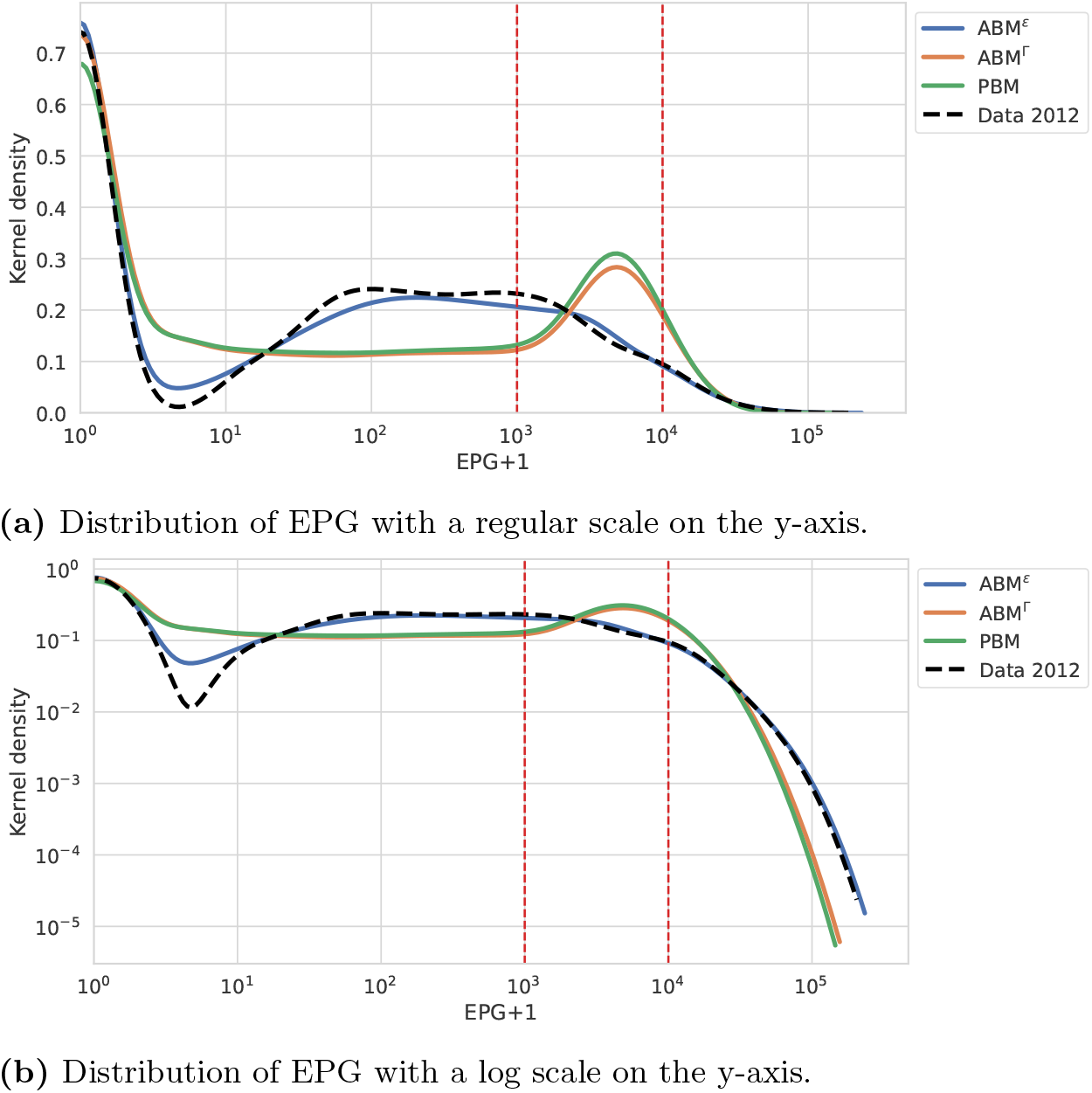
Kernel density estimate of EPG in the 2012 data and in the equilibrium state of all models on a regular and on a log scale on the y-axis. The x-axis is plotted on a log scale. The dashed vertical red lines indicate the transition from low to medium and from medium to high EPG burden, respectively. The PBM and the ABM^Γ^ show very strong overlap.

The goodness of fit can also be evaluated by comparing the 2012 data with the model states in equilibrium at the beginning of the time series in Fig 5. While all models can fit the prevalence and mean worm burden targets (Fig 5a), only the ABM^*ε*^ can fit all distributional targets as well (Fig 5b). The PBM and the ABM^Γ^ show the same pattern in derivation from the data at equilibrium: They underestimate the prevalence and mean EPG for the group with low worm burden while the reverse is true for the moderate worm burden: both the PBM and the ABM^Γ^ overestimate the prevalence of individuals in this group and the mean EPG within this group. Finally, for the high worm burden, there is a slight underestimation of the mean worm burden within this group by the ABM^Γ^ and the PBM, while all models fit the prevalence of high worm burden well.

**Fig 5.**
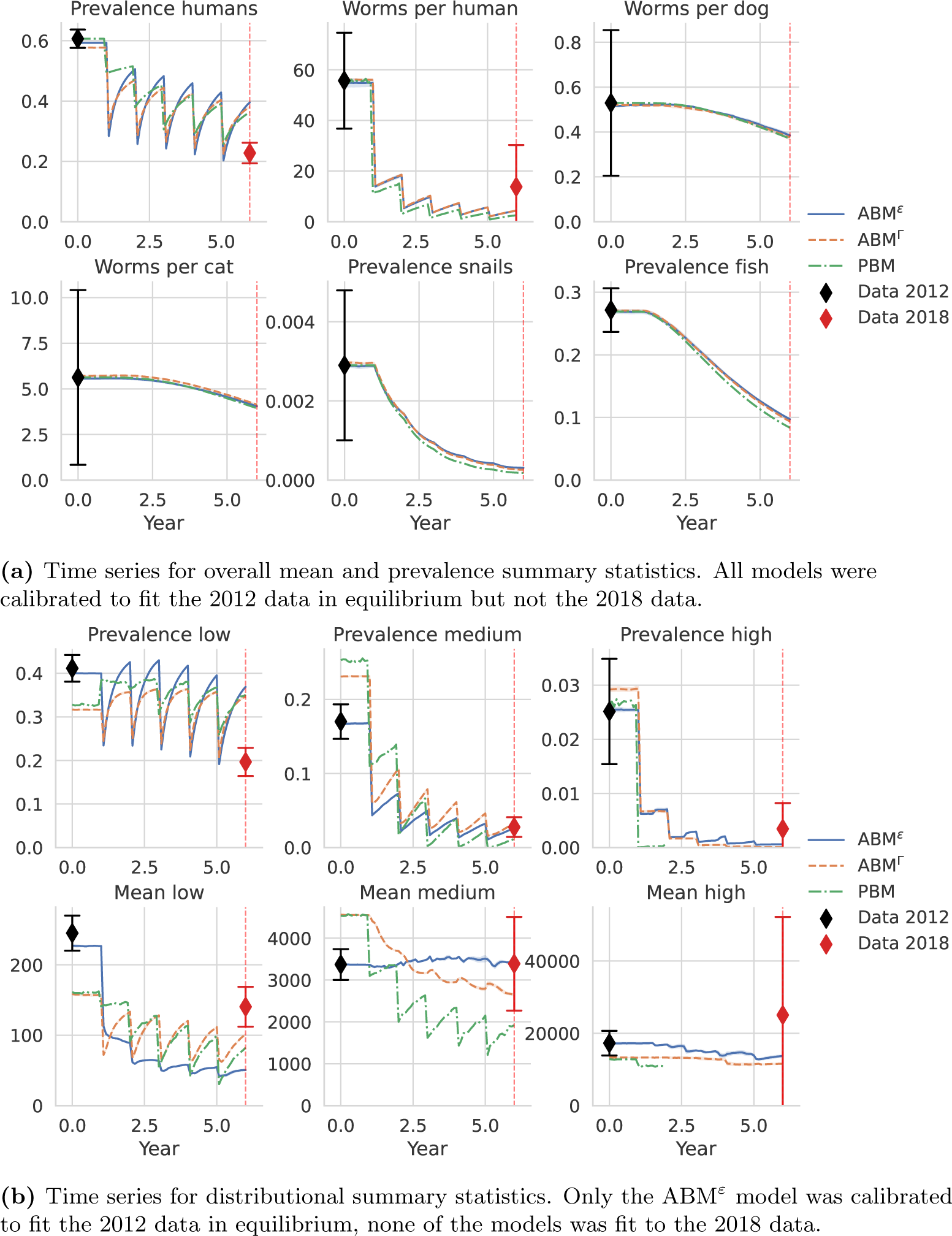
Time series showing the mean of the summary statistics from model runs with 10 random seeds for each model variant. The error bands indicate the minimum and maximum values over the random seeds. All runs start at the fitted equilibrium. After 1 year in equilibrium, yearly MDA is conducted at a coverage of 80%.

Fig 5 also allows the comparison of the impact prediction of the MDA campaign with 80% coverage made by the models. There is a remarkable agreement between the predictions of all models with regards to the prevalence and overall mean worm burden variables (Fig 5a). The most notable difference is the smaller drop in prevalence predicted by the PBM after the initial distributions of MDA.

There is more difference between the predictions of the models in terms of distributional summary statistics (Fig 5b). Most notably, the PBM predicts a sharp drop in the number of individuals with high worm burden with no more individuals remaining in this category after 2 years of MDA campaign. While the predictions of the models mostly agree in terms of prevalence of low and medium worm burden, the mean EPG within these groups shows some disagreement.

Comparing the predictions of the models in Fig 5 with field data measured after 5 rounds of yearly MDA (“Data 2018”), we see that all models overestimate the remaining prevalence by a factor of almost 2 (Fig 5a). All models underestimate worms per human, though the predictions still lie within the confidence interval, which is large for this variable due to the high degree of aggregation. From Fig 5b, we see that the overestimation in prevalence is entirely attributable to an overestimation of the prevalence of individuals with a low worm burden, while the prediction of moderate worm burden prevalence matches the data well and the predicted high worm burden prevalence lies below the data though within the confidence interval.

Fig 6 depicts the value of the state variables after 5 years of yearly MDA (indicated by the red dashed vertical lines in Fig 5) for coverages between 0% and 90%. The high agreement between models for the overall mean and prevalence variables persists over the entire range of coverages (Fig 6a). For the distributional summary statistics, there is more disagreement between the models over the range of coverages, again with the ABM^Γ^ and the PBM being closer together in predictions (Fig 6b). Fig 6 again includes the 2018 data. We see that no model predicts a reduction prevalence as seen in the data with any level of coverage between 0% and 90%. On the other hand, all models predict the reduction in mean worm burden to match the field data at a coverage of 40%, which lies below the estimated coverage 80% under which the 2018 data was generated. For the distributional summary statistics, the 2018 data can be reproduced for all variables except for the overall prevalence, the prevalence of low mean worm burden and the mean EPG in the group of high worm burden. However, the required coverage for attaining the targets varies over summary statistics.

**Fig 6.**
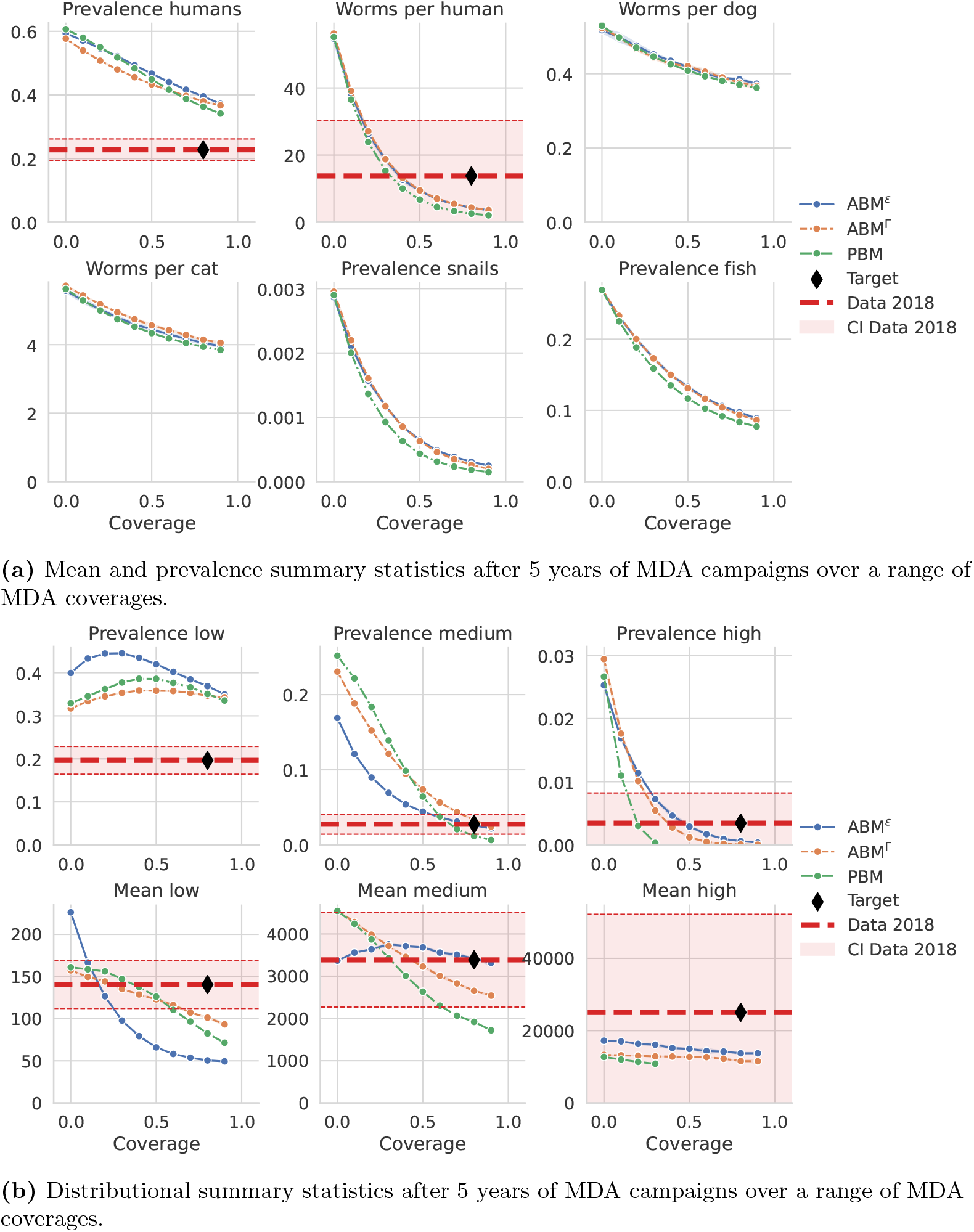
Summary statistics after 5 years of yearly MDA campaign at the time point indicated by the vertical red dashed line in Fig 5. Each model variant is run with 10 random seeds for each MDA coverage and the mean is taken for the respective summary statistic. The error bands show the minimum and maximum values observed over the random seeds. The data from the year 2018 is plotted if available. At an MDA coverage of 80%, the model prediction should ideally match the data, as indicated by the black diamond.

Fig 7 shows which combinations of education campaign efficacy and variance in the beta distribution of MDA participation (“MDA variance”) are required in the model under the assumption of an 80% MDA coverage to achieve the same prevalence and mean worm burden as seen in the 2018 data. Both target prevalence and target mean worm burden are achieved at the intersection of the lines which is indicated by the marker labeled “optimum”. The ABM^Γ^ requires an MDA variance of 0.24 and an education efficacy of 45% while the ABM^*ε*^ requires a lower MDA variance of 0.21 as well as a lower education efficacy of 33%. Fig 7 also indicates which change to the intervention is important for replicating which variable. For both model variants, the target prevalence can be achieved with a wide range of MDA variance levels while the education efficacy is required to lie in a much narrower band. Therefore, the model predicts the education campaign to be main driver for the observed reduction in prevalence. Conversely, the heterogeneity in MDA adherence is the main driver of the observed persistence of substantial mean worm burden across the population. Fig 8 shows model runs using the optimal values of MDA variance and education campaign efficacy as indicated in Fig 7. The prevalence and mean worm burden now fit the 2018 data as expected. The distributional summary statistics in Fig 8b show that the reduction in prevalence is mainly achieved by reducing the prevalence of individuals with low worm burden. In fact, the model prediction of low worm burden prevalence now fits the 2018 data even though it was not used to fit the adapted intervention characteristics. The increase in mean worm burden by MDA variance is mainly achieved through increased prevalence of high mean worm burden. While the models without MDA variance estimated very little persistence of high worm burden, the models with MDA variance predict high worm burden at the upper end or above the confidence interval of the 2018 data.

**Fig 7.**
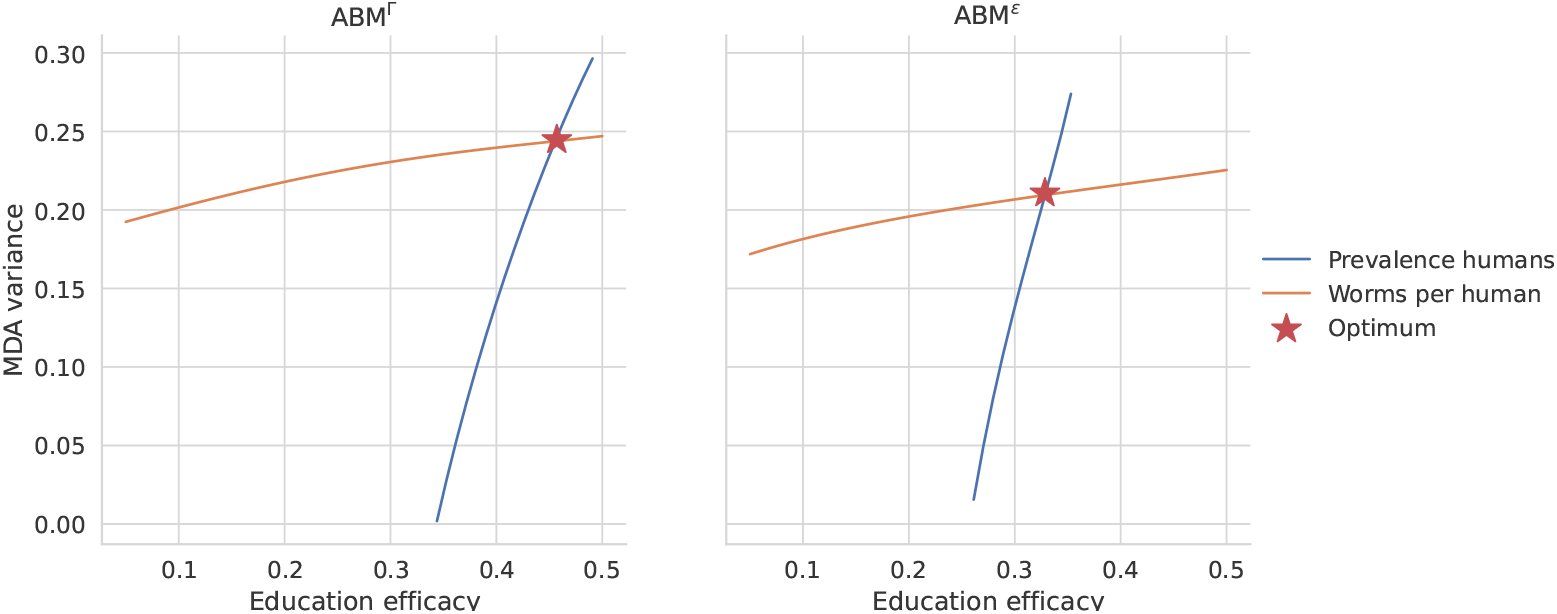
Required combinations of education campaign efficacy and MDA variance in the agent-based models to attain the same prevalence and intensity of infection as seen in the 2018 data after 5 MDA campaigns with an estimated coverage of 80%. Both target values are met at the intersection of both lines, as indicated by the marker labeled “optimum”.

**Fig 8.**
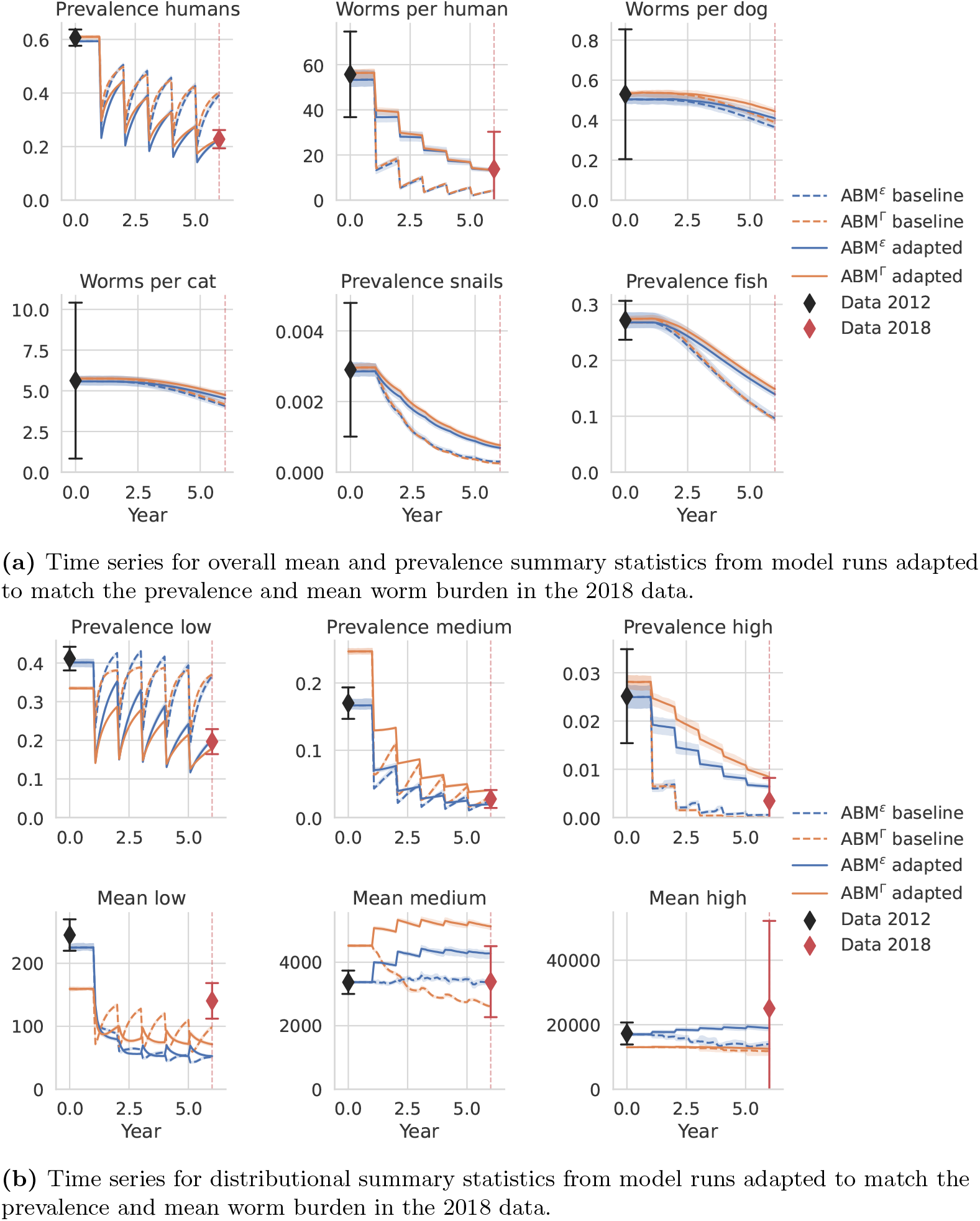
Simulations run with MDA variance and education efficacy such that target reduction in mean worm burden and prevalence can be achieved. The predictions for mean worm burden and prevalence now match the 2018 data. The “baseline” model runs without the adaptations are also plotted for comparison and are the same as in Fig 5.

Fig 9 depicts the evolution of the aggregation parameter *k* for worms over time under the MDA campaigns with 80% coverage in the PBM as well as the ABMs with and without fitted education campaigns and heterogeneity in MDA adherence. The field data which is also plotted shows a decrease in the aggregation parameter *k* when comparing the data from before the MDA campaigns in 2012 to the data collected after MDA campaigns in 2018. Low prevalence after interventions has also been associated with a high degree of aggregation for hookworm infection [31]. For the PBM, *k* is constant by assumption. For the ABMs without education and heterogeneity in MDA adherence, *k* drops sharply after each round of MDA. This indicates an increased degree of heterogeneity which occurs because individuals who received MDA are now free of worms and the remaining worm burden is carried only by those 20% of individuals that did not receive MDA. As time passes since the last round of MDA, individuals that received MDA start acquiring worms again and catch up in worm counts compared to those who did not receive MDA which reduces aggregation and therefore increases *k*. The ABMs with the adapted heterogeneity in MDA adherence and education campaign exhibit sustained reductions in *k*. This is because individuals with lowest worm uptake rates are most likely to be reached by the MDA campaign. These individuals are slower to start taking up worms again after MDA, which would increase prevalence and therefore *k*. In addition, the education campaigns further reduce the uptake rates of all individuals and therefore slow down the rebound of prevalence after an MDA campaign. The prediction of *k* perfectly fits the 2018 data for the ABMs with education campaigns and the heterogeneity in MDA adherence because the MDA variance and education campaign efficacy were chosen to fit the mean worm burden and the prevalence, which are the sole determinants of the aggregation parameter *k*.

**Fig 9.**
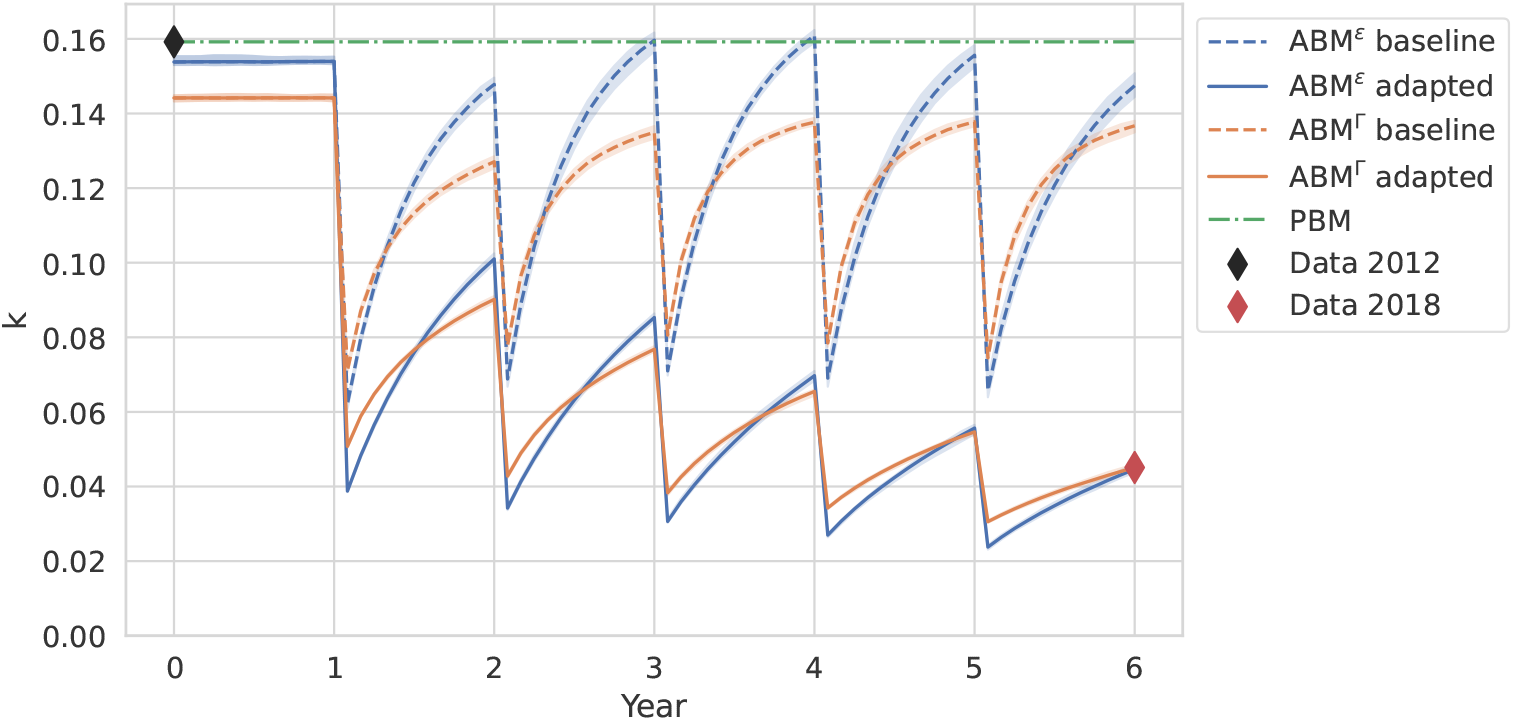
Time series of aggregation parameter *k* for worms in indivudals in the model runs depicted in Fig 8. For the ABMs, values are calculated by numerically solving Equation (3) for *k* given the mean worm burden and prevalence at each time step. For the PBM, *k* is constant over time by assumption.

## 4 Discussion

Previous PBMs of *Opisthorchis viverrini* could accurately capture the prevalence of infection in reservoir and intermediate hosts, as well as the prevalence of infection and mean worm burden in humans [15, 25]. However, as compartmental models, they could not capture the distribution of worm burden in humans, nor how the relationship between prevalence of infection and mean worm burden changes due to repeated MDA campaigns. Here, we presented the first AMBs of *O. viverrini* transmission that allow us to incorporate heterogeneities among humans in uptake of the parasite and adherence to treatment in order to better reproduce the distribution of worm burden in humans before and after mass-drug administration campaigns.

We conducted a thorough comparison of the ABMs with the PBM to show the utility of both types of models. Both perform similarly well in reproducing the impact of interventions in transmission reduction, again emphasizing the utility of PBMs in macroparasite modelling. However, the ABMs are better able to capture the distribution changes in response to MDA campaigns when incorporating plausible heterogeneities in MDA adherence combined with education campaigns.

We show that education campaigns, and therefore behavior change, was necessary to explain the reduction in prevalence; and that this could not be explained solely by MDA campaigns. On the other hand, we show that a high degree of heterogeneity in MDA adherence is a plausible cause for the persistence of high worm burden in some individuals, highlighting the importance of reaching these individuals in future MDA campaigns.

The mechanisms of systematic MDA adherence and an education campaign can be easily added to ABMs. For the compartmental model, in contrast, modelling an evolution of the aggregation parameter *k* would become necessary and it is not clear how heterogeneity in MDA adherence and an education campaign would translate to changes of *k* (see also Fig 9). In contrast to results from ABMs for hookworm epidemiology and control [32], we find that the aggregation parameter *k* is not increasingly reduced over multiple yearly MDA campaigns when assuming random adherence due to quick rebounds in infection rates.

There are several important factors we have not considered for simplicity or because of data unavailability. MDA campaigns have been conducted in the study area prior to 2012, albeit on an irregular basis [11] and we lack data on the timing and the coverage of these campaigns. The assumption of an equilibrium at the start of the simulation period may therefore be inappropriate and yield parameter fits which underestimate transmission intensities. However, relaxing this assumption would require additional data on the timing and coverage levels of these interventions.

We have also not considered inaccuracies of the EPG measurement technique. Specifically, sensitivity may not be perfect, leading to an underestimation of prevalence, and the EPG counts for high worm burden may be subject to large variances. We also assumed MDA to have perfect efficacy, leading to clearance of all parasites when an individual receives the drug. This assumption is not realistic, especially in individuals with high worm burden, but there is little data to better estimate the efficacy of the campaigns. Even though we have sample sizes of several hundred individuals, the uncertainty for the proportion of high worm burden individuals is high as only very few individuals fall into this category. Specifically, for the year 2012 there were 25 individuals with high worm burden, while in 2018, there were only two individuals. In the absence of data that could provide realistic assumptions of EPG measurement sensitivity or MDA efficacy, we chose the simplifying but extreme assumption of perfect diagnostic sensitivity and perfect MDA efficacy.

An obvious drawback of the ABM^*ε*^ is the direct derivation of the transmission parameter distribution from the field data, which means that we are using the data we fit to also to parameterize the model. While this may limit the generalizability of results, it may be appropriate for studying a specific population. There may also be situations where the field data cannot be captured as well with a gamma distribution, and the results suggest that ABM^*ε*^ may be a valid alternative in such a situation. One could also criticize that ABM^*ε*^ fixes the proportion of people with a worm uptake rate equal to zero and thereby the prevalence at equilibrium. However, the ABM^Γ^ effectively does the same by drawing a low value of *β*_*hf,i*_ for some individuals such that they never have any parasite uptake. While the ABMs allow setting a lot of parameters at the level of individuals, we often lack information on how to specify these variables. For example, the choice of a beta distribution for the MDA adherence was chosen for simplicity and may not reflect reality. We also did not include heterogeneity in the efficacy of the education campaign both, for simplicity and due to a lack of data, even though heterogeneity is likely to exist in reality. We expect that including heterogeneity in education campaign efficacy at the individual level will add a degree of freedom that would allow us to even better model the impact of the interventions on the distribution of worm burden in the population.

A clear next step is to integrate the ABMs with disease models as they better capture individuals with the highest worm burden and therefore the highest risk of acquiring chronic disease. Furthermore ABMs allow us to track the treatment history and the aggregate worm burden over time of each individual, as well as additional risk factors. Currently ongoing studies associating worm burden and disease will allow the appropriate parameterization of such models that can then be used to predict the impact of interventions on the incidence of cholangiocarcinoma based on the history of infection and treatment at an individual level.

## Data Availability

The model and data will be made publicly available on a github repository.

## Supporting information

- Supplementary information: Detailed model descriptions of ABMs and PBM

## Notes

### Competing Interest Statement

The authors have declared no competing interest.

